# Why Invariant Risk Minimization Fails on Tabular Data: A Gradient Variance Solution

**DOI:** 10.64898/2026.04.09.26350513

**Authors:** Grold Otieno Mboya

**Affiliations:** School of Health Sciences, Department of Epidemiology and Biostatistics Jaramogi Oginga Odinga University of Science and Technology, Kenya

**Keywords:** distribution shift, invariant learning, causal inference, tabular data, gradient variance regularization, calibration, epidemiology

## Abstract

Machine learning models trained on observational data from one environment frequently fail when deployed in another, because standard learning algorithms exploit spurious correlations alongside causal ones. Invariant learning methods address this problem by seeking representations that support stable prediction across training environments, but their behavior on tabular data remains poorly characterized. We present CausTab, a gradient variance regularization framework for causal invariant representation learning on mixed tabular data. CausTab penalizes the variance of parameter gradients across training environments, providing a richer invariance signal than the scalar penalty used by Invariant Risk Minimization (IRM). We provide formal results showing that the gradient variance penalty is zero at causally invariant solutions and positive at solutions that rely on spurious features. Through experiments on synthetic data across three spurious-correlation regimes, four cycles of the National Health and Nutrition Examination Survey (NHANES), and four hospital systems in the UCI Heart Disease dataset, we demonstrate that: (1) IRM consistently degrades relative to standard empirical risk minimization (ERM) on tabular data, losing up to 13.8 AUC points in spurious-dominant settings, a failure we trace mechanistically to penalty collapse during training; (2) CausTab matches or exceeds ERM in every experimental condition; (3) CausTab achieves consistently better probability calibration than both ERM and IRM; and (4) invariant learning methods fail when environments differ in outcome prevalence rather than in spurious feature correlations, a boundary condition we characterize both empirically and theoretically. We introduce the Spurious Dominance Index (SDI), a practical scalar diagnostic for determining whether a dataset requires invariant learning, and validate it across all experimental settings.

## 1 Introduction

A predictive model that performs well on training data can fail substantially when deployed in a different context. This is not a subtle statistical phenomenon. Models trained on data from one hospital system can degrade when applied to another. Models trained on survey data from one decade perform differently on data collected a decade later. Models built on populations from one country may not transfer to populations with different demographic compositions. The failure mode is well-understood: standard learning algorithms optimize for average performance on the training distribution and exploit all available statistical associations, including those that are incidental to the specific context in which the training data were collected. When those incidental associations shift at deployment, predictions degrade. This problem is known as distribution shift, and it is one of the most common causes of real-world model failure in healthcare, epidemiology, and public policy (Ben-David et al., 2010; Saria and Subbaswamy, 2019).

The causal lens makes this failure precise. Statistical associations in observational data arise from two distinct sources. Causal relationships arise when a feature directly influences the outcome through the data-generating mechanism: blood pressure causally affects cardiovascular risk; age causally affects hypertension onset. These relationships are stable across environments because the underlying biological mechanism does not change when we move from one hospital to another or from one survey year to the next. Spurious correlations, by contrast, arise when a feature is associated with the outcome only through confounding variables or environment-specific selection mechanisms. A demographic variable that correlates with hypertension in a given survey cycle because of healthcare access patterns in that cycle may not carry the same correlation when access patterns change. A model that learns to rely on spurious correlations will perform well where those correlations hold and degrade wherever they do not.

Invariant learning methods aim to exploit this distinction by finding representations that support equally accurate prediction across all training environments. The theoretical foundation was established by Peters et al. (2016) through invariant causal prediction, which identified causal features as precisely those for which the conditional distribution *P* (*y* | **x**^*c*^) is stable across environments. Arjovsky et al. (2019) operationalized this idea for neural networks through Invariant Risk Minimization (IRM), which encourages the learned representation to support an invariant linear classifier across training environments. Despite this theoretical appeal, however, these methods have not consistently outperformed standard empirical risk minimization (ERM) in practice. Gulrajani and Lopez-Paz (2021) demonstrate that IRM frequently underperforms ERM on the DomainBed benchmark, and Rosenfeld et al. (2021) identify theoretical conditions under which IRM fails to recover invariant solutions.

Critically, both of these analyses focus on image classification tasks. The behavior of invariant learning methods on tabular data, which is the dominant format in healthcare, epidemiology, economics, and social science, has not been systematically studied. This is a consequential gap. The practitioners who most urgently need distribution-shift-robust methods are not building image classifiers; they are building clinical risk scores, epidemiological prediction models, and policy evaluation tools, all of which operate on tabular data. We address this gap directly. We make four contributions.

### CausTab

We present a gradient variance regularization framework for invariant learning on tabular data. CausTab penalizes the variance of parameter gradients across training environments. A parameter that responds to a causal feature receives a consistent gradient signal across environments because the feature contributes similarly to the loss wherever the data came from. A parameter that responds to a spurious feature receives an inconsistent signal because the spurious correlation varies by environment. Penalizing gradient variance therefore discourages reliance on spurious features without penalizing reliance on causal ones. Unlike IRM, which uses a scalar dummy variable to probe invariance at the output layer only, CausTab uses the full gradient vector across all parameters, providing a richer signal that is harder to satisfy through superficial adjustments.

### IRM failure analysis

We provide the first systematic empirical documentation of IRM’s failure on tabular data. Across three synthetic spurious-correlation regimes and a continuous spurious strength sweep spanning seven levels, IRM consistently underperforms ERM by up to 13.8 AUC points. We trace this failure mechanistically to penalty collapse: IRM’s scalar constraint declines toward zero during training, ceasing to enforce invariance. CausTab does not exhibit this behavior.

### Spurious Dominance Index

We introduce SDI, a scalar diagnostic computed from training data that characterizes whether a dataset is in a regime where invariant learning is likely to help. We validate SDI across three synthetic regimes and two real datasets with fundamentally different types of distribution shift.

### Calibration analysis

We conduct the first calibration analysis of invariant learning methods on tabular data. CausTab achieves consistently lower expected calibration error (ECE) than ERM and IRM across all NHANES experiments. In clinical settings where stated probabilities directly influence treatment decisions, this calibration advantage is practically significant independently of AUC.

We also document and explain a boundary condition: invariant learning methods, including CausTab, fail when environments differ in outcome prevalence rather than in spurious feature correlations. This occurs because the shared causal mechanism assumption that underlies all invariant learning is violated. We characterize this failure empirically on the UCI Heart Disease dataset and discuss practical implications for practitioners who may encounter similar settings.

The remainder of this paper is organized as follows. Section 2 reviews related work. Section 3 formalizes the problem. Section 4 presents the CausTab framework. Section 5 provides theoretical analysis. Section 6 describes experiments and results. Section 7 discusses findings and limitations. Section 8 concludes.

## 2 Related Work

### Invariant causal prediction

The theoretical foundation for invariant learning was established by Peters et al. (2016), who formalized the idea that causal features are precisely those for which the conditional distribution *P* (*y* | **x**^*c*^) is stable across environments. They proved consistency of a testing procedure for identifying such features under linear structural equation models and provided a framework for constructing confidence intervals for causal effects. Their work established the key insight that stability of predictive relationships across heterogeneous settings is both necessary and sufficient for causal identification under certain structural assumptions.

### Invariant risk minimization

Arjovsky et al. (2019) operationalized the invariant prediction principle for neural networks through the IRM penalty, which encourages a learned representation to support an invariant linear classifier across all training environments. Their theoretical analysis proves that IRM recovers the causal invariant predictor under linear models with sufficient environments. Subsequent work has identified important limitations. Rosenfeld et al. (2021) show that IRM can fail when training environments are insufficiently diverse and that the required number of environments grows with the dimensionality of the spurious feature space. Gulrajani and Lopez-Paz (2021) demonstrate empirically that IRM frequently underperforms ERM on the DomainBed benchmark suite, raising questions about its practical utility. Ahuja et al. (2021) connect IRM to the information bottleneck principle and provide tighter conditions under which invariant learning succeeds. Krueger et al. (2021) propose Risk Extrapolation (V-REx), which penalizes variance of per-environment losses rather than gradient norms, and prove stronger guarantees in some linear settings. Our work is related to V-REx in spirit but differs in operating at the gradient level: penalizing gradient variance is sensitive to individual parameter behavior and harder to circumvent through output-layer adjustments.

### Domain generalization

Domain generalization methods seek predictors that perform well on unseen target domains given data from multiple source domains during training. Distribution alignment approaches reduce discrepancy between source and target feature distributions using kernel-based statistics such as maximum mean discrepancy (Gretton et al., 2012). These methods do not distinguish causal from spurious features and can erase useful causal signal along with the spurious noise when the two are correlated. Distributionally robust optimization (Sagawa et al., 2020) optimizes for worst-case performance over an uncertainty set of distributions, providing minimax guarantees without requiring environment labels. This is conservative by design and may sacrifice average-case performance. Our experimental evaluation focuses on IRM as the most directly comparable invariant learning baseline, since it shares the same theoretical motivation as CausTab.

### Causal representation learning

Schölkopf et al. (2021) provide a comprehensive framework for causal representation learning, arguing that robust generalization requires models that respect the causal structure of the data-generating process. They distinguish between interventional and observational robustness and connect representation learning to structural causal models. CausTab is a practical instantiation of this principle for tabular data, making no assumptions about the causal graph while enforcing the observable implication of causal invariance: stable gradient signals across environments.

### Distribution shift in healthcare

Distribution shift in clinical and epidemiological prediction models has been documented across hospital systems (Subbaswamy et al., 2020), geographic regions, and time periods. The epidemiological context is particularly relevant because temporal distribution shift in survey data is both common and consequential: demographic compositions change, disease management guidelines evolve, and measurement protocols are updated. Standard operational responses include periodic model retraining, threshold recalibration, and deployment monitoring. The present work contributes a principled training objective for producing more robust models at the outset, complementing these operational strategies.

### Tabular deep learning

Deep learning models for tabular data have received renewed attention following the observation that carefully designed architectures and training procedures can approach the performance of gradient boosted trees on some benchmarks (Gorishniy et al., 2021). Our work does not propose a new architecture. We use a standard feedforward network and focus entirely on the training objective, ensuring that observed performance differences across methods are attributable to the invariance penalty and not to architectural choices.

## 3 Problem Formulation

### 3.1 Setting

Let **x** ∈ 𝒳 ⊆ ℝ^*d*^ denote a *d*-dimensional feature vector and *y* ∈ {0, 1} a binary outcome. Data are collected across *E* distinct environments ℰ = {*e*_1_, …, *e*_*E*_}, where each environment *e* yields an independent dataset

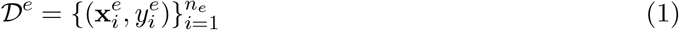

drawn from a joint distribution *P*^*e*^(**x**, *y*). We make no assumption that *P*^*e*^ is identical across environments. In the applications studied in this paper, environments correspond to distinct time periods (NHANES survey cycles) or distinct institutional settings (hospital systems in the UCI Heart Disease dataset), and the distributional differences between them constitute the distribution shift we seek to address.

### 3.2 Causal and Spurious Features

We partition the feature vector into two conceptually distinct components:

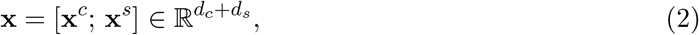

where 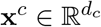 are *causal features* whose relationship with *y* is invariant across environments, and 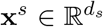 are *spurious features* whose relationship with *y* varies across environments due to confounding, selection bias, or measurement shift. This partition is a conceptual device: in practice it is unknown, and recovering it from observational data is precisely the goal of invariant learning.

#### Definition 1

(Causal invariance). *A feature subset* **x**^*c*^ *satisfies* causal invariance *if*

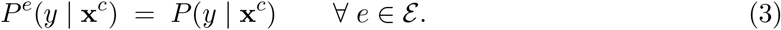

This definition states that **x**^*c*^ carries the same information about *y* regardless of which environment the data came from. Spurious features violate this condition: there exist environments *e, e*′ ∈ ℰ such that *P*^*e*^(*y* | **x**^*s*^) ≠ *P*^*e′*^ (*y* | **x**^*s*^). The predictive relationship between a spurious feature and the outcome is environment-dependent and will shift whenever the data-collection context changes.

### 3.3 The Distribution Shift Problem

Standard empirical risk minimization (ERM) learns a predictor *f*_***θ***: 𝒳→[0,1]_ by minimizing average prediction error across all training environments:

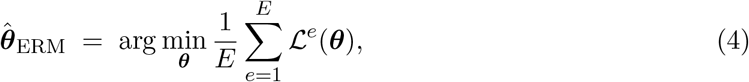

where 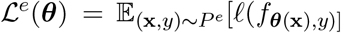 and ℓ is the binary cross-entropy loss. ERM treats all statistical associations identically, making no distinction between causal and spurious signal. When spurious correlations shift between training and deployment, the ERM predictor degrades in proportion to its reliance on those correlations.

**Remark 1** (Implicit ERM robustness). *ERM can achieve approximate robustness by accident when causal features dominate the predictive signal. If a single strong causal predictor outweighs all spurious features in terms of predictive power, ERM gravitates toward relying on the causal feature not because it identifies it as causal, but because it is the most predictive signal available. Our NHANES experiments demonstrate this phenomenon empirically, and the Spurious Dominance Index formalizes when it can be expected to occur*.

### 3.4 The Invariant Learning Objective

We seek a representation *ϕ*: 𝒳 → *Ƶ* and predictor *g*: *Ƶ* → [0, 1] satisfying two conditions simultaneously:

1. **Predictive performance**. The composed predictor *g* ◦ *ϕ* achieves low cross-entropy loss across all environments.
2. **Invariance**. The conditional distribution *P*^*e*^(*y* | *ϕ*(**x**)) is identical across all *e* ∈ ℰ.

Condition 1 ensures the representation is useful for prediction. Condition 2 ensures it has removed environment-specific spurious signal, so that the information retained in *Ƶ* reflects the causal relationship between features and outcome. The challenge is enforcing Condition 2 without access to the true causal graph, using only observational data from multiple environments.

## 4 The CausTab Framework

### 4.1 Architecture

CausTab uses a feedforward neural network 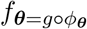, where *ϕ*_***θ***:*X*→*Ƶ*_ is an encoder mapping inputs to a *k*-dimensional representation and *g*: *Ƶ* → [0, 1] is a linear classification head followed by a sigmoid activation. The encoder consists of two fully connected layers with batch normalization, ReLU activations, and dropout regularization. All three methods compared in this paper (ERM, IRM, CausTab) use the same architecture with hidden widths 128 and 64 respectively. This design choice is deliberate: it ensures that any observed performance difference across methods is attributable to the training objective and not to differences in model capacity or inductive bias.

### 4.2 The Gradient Variance Penalty

For each environment *e* ∈ ℰ, define the environment-specific gradient vector as

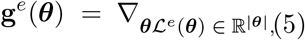

where |***θ***| denotes the total number of parameters. The gradient variance penalty is then

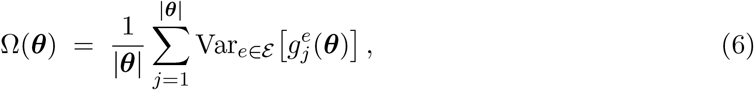

where 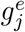 denotes the *j*-th component of **g**^*e*^(***θ***) and Var_*e*∈ℰ_ [·] denotes variance computed across the *E* environments. The division by |***θ***| normalizes the penalty to be independent of model size, which makes the penalty weight *λ* interpretable and comparable across architectures.

The full CausTab training objective is

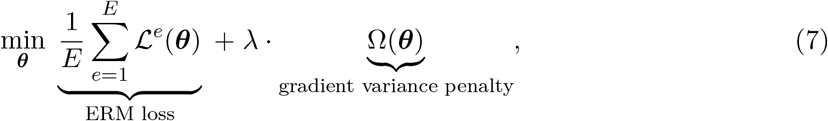

where *λ >* 0 controls the relative weight of the invariance penalty.

#### Intuition

Consider a parameter *j* that predominantly responds to a causal feature. Because the causal feature contributes similarly to the loss regardless of the environment, the gradient 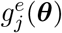 is consistent across environments, and its variance is small. Now consider a parameter that predominantly responds to a spurious feature. Because the spurious correlation varies by environment, the gradient it receives varies accordingly, and its variance is large. The penalty Ω(***θ***) aggregates these per-parameter variances and adds the sum to the loss, discouraging the network from building representations that depend on environment-sensitive parameters.

### 4.3 Comparison with IRM

Arjovsky et al. (2019) enforce invariance through a scalar penalty applied per environment:

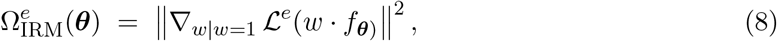

where *w* ∈ ℝ is a scalar dummy variable. This penalty measures whether a scalar rescaling of the predictions can reduce the loss in environment *e* and penalizes any such rescaling.

CausTab differs from IRM in two important respects. First, CausTab uses the full gradient vector **g**^*e*^ ∈ ℝ^|***θ***|^ rather than a scalar dummy gradient. This is a strictly richer signal: the full gradient captures how each individual parameter responds to each environment, whereas IRM’s scalar captures only an aggregate response at the output layer. Second, CausTab penalizes the variance of gradients across environments, not the magnitude of a per-environment gradient. A parameter with a consistently large gradient across all environments is not penalized by CausTab, because consistency indicates causal relevance. IRM would penalize such a parameter if its per-environment gradient is nonzero.

These differences have direct practical consequences. IRM’s scalar penalty can be satisfied by small adjustments to the output layer weights without any change to the internal representation, because the dummy variable *w* is applied only at the output. This is the mechanism underlying the penalty collapse we document in Section 6. CausTab’s full gradient penalty cannot be satisfied in this way: it requires the gradient signal at every parameter throughout the network to be consistent across environments.

### 4.4 Training Procedure

Training proceeds in two phases controlled by an annealing and warmup schedule.

#### Phase 1: ERM warmup (epochs 1 to *T*_*a*_)

For the first *T*_*a*_ epochs, CausTab trains with *λ* = 0, equivalent to standard ERM. This allows the network to develop a reasonable predictive solution before invariance pressure is applied. Applying the full penalty from the first epoch is counterproductive because the initial random parameters have no meaningful relationship to any feature, causal or spurious, and the resulting gradients carry no informative signal about causal structure.

#### Phase 2: Invariance enforcement (epochs *T*_*a*_ + 1 to *T*)

After epoch *T*_*a*_, the penalty weight ramps linearly from 0 to *λ* over *T*_*w*_ epochs:

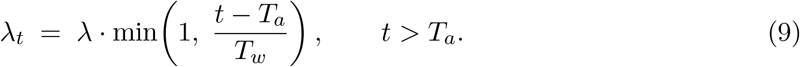

This linear warmup prevents a sudden large penalty from destabilizing the optimization at epoch *T*_*a*_. The ablation study in Section 6.5 confirms that omitting the warmup ramp produces slightly less stable results, though the differences are small in the settings we study.

In all experiments we use *T*_*a*_ = 50, *T*_*w*_ = 20, *T* = 200, *λ* = 100, and optimize with Adam (Kingma and Ba, 2015) at learning rate *η* = 10^−3^. The complete training procedure is given in Algorithm 1.

##### Algorithm 1

CausTab Training Algorithm

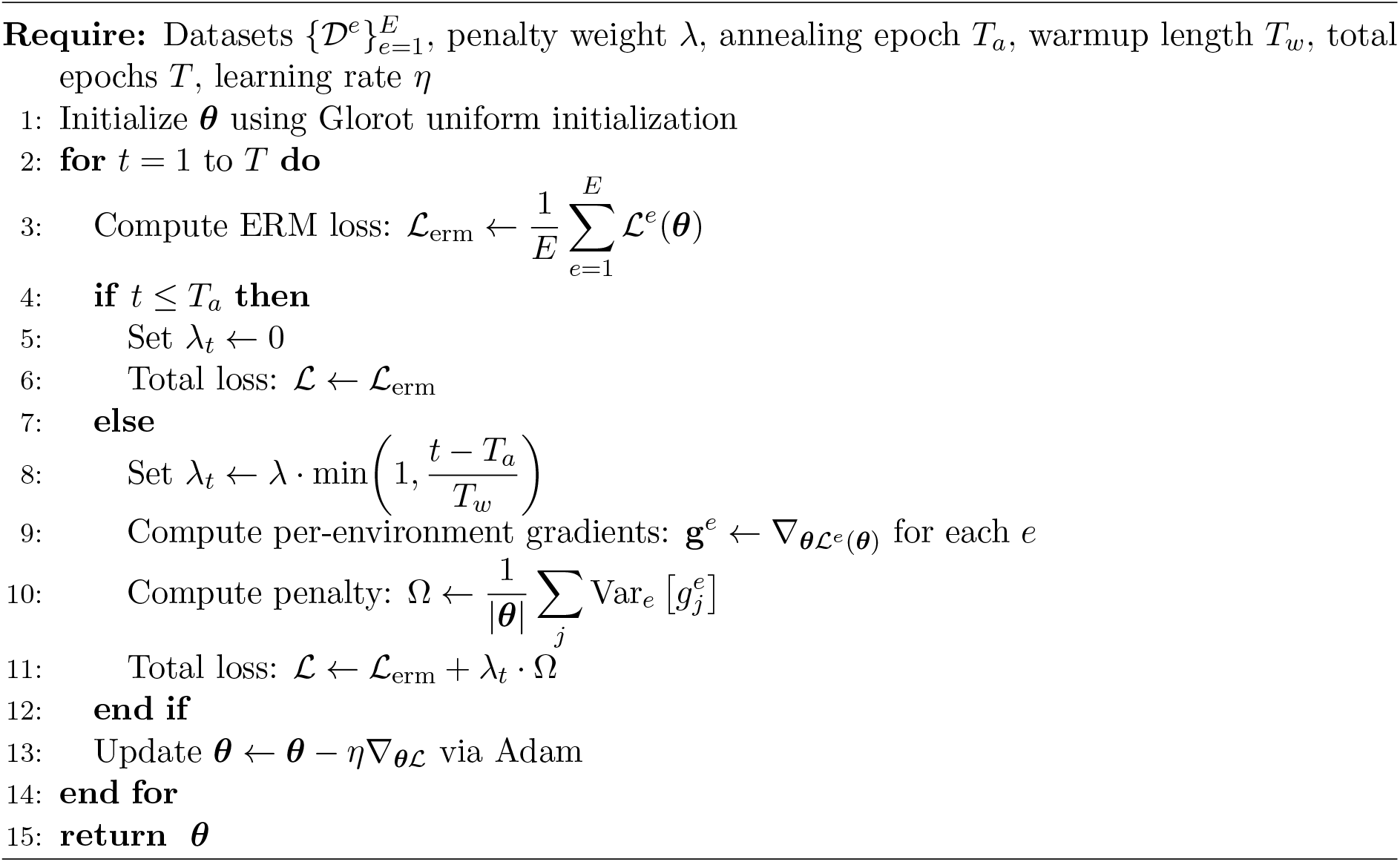

### 4.5 The Spurious Dominance Index

A practitioner considering whether to use CausTab faces a natural question: does the target dataset actually exhibit the kind of distribution shift that invariant learning is designed to address? We introduce the Spurious Dominance Index (SDI) as a practical diagnostic computed from the training data alone.

#### Definition 2

(Spurious Dominance Index). *Let* 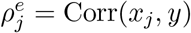 *denote the Pearson correlation of feature j with the outcome in environment e. Define the stability of feature j across environments as*

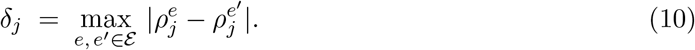

*Classify feature j as spurious if* 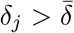, *the median stability across all features, and as causal otherwise. Let* 𝒮 *and* 𝒞 *denote the resulting sets. The Spurious Dominance Index is*

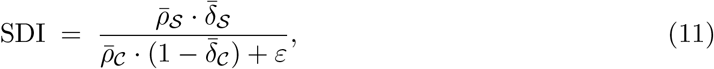

*where* 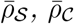 *denote mean absolute correlations within each set*, 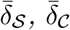 *denote mean stabilities, and ε >* 0 *prevents division by zero*.

SDI is large when features that are strongly predictive of the outcome are also unstable across environments, indicating that spurious correlations dominate the predictive signal. SDI is small when the strongest predictors are stable, indicating that ERM is likely to be implicitly robust. We validate SDI empirically in Section 6.7.

## 5 Theoretical Analysis

We ground CausTab in theory by identifying conditions under which the gradient variance penalty has the correct directional properties. We state each assumption explicitly and discuss its practical implications.

### Assumption 1

(Sufficient environments). *For every spurious feature j* ∈ 𝒮, *there exist environments e, e*′ ∈ ℰ *such that* 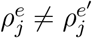.

### Assumption 2

(Shared causal mechanism). *The conditional distribution P*^*e*^(*y* | **x**^*c*^) *is identical across all environments e* ∈ ℰ.

### Assumption 3

(Realizability). *There exists* ***θ***^∗^ ∈ ℝ^|***θ***|^ *such that* 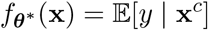.

Assumption 1 requires that training environments are diverse with respect to spurious features. It fails when environments are too similar, which explains why all three methods perform comparably on NHANES data where temporal shift is moderate. Assumption 2 is the structural assumption that underlies all invariant learning: the data-generating mechanism for *y* given the causal features is the same across environments. It fails when environments differ in the causal mechanism itself rather than in their confounding structure, as we document for the UCI Heart Disease dataset in Section 6.4. Assumption 3 is standard in learning theory and requires that the model class is expressive enough to represent the true causal predictor.

### Theorem 1

(Gradient variance at the causal solution). *Under Assumptions 1–3, the parameter vector* ***θ***^∗^ *satisfying causal invariance (Definition 1*) *achieves* Ω(***θ***^∗^) = 0.

*Proof*. At ***θ***^∗^, by assumption we have 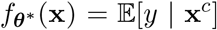. Since *P*^*e*^(*y* | **x**^*c*^) is identical across environments (Assumption 2), the loss ℒ^*e*^(***θ***^∗^) is identical across environments. It follows that 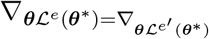 for all *e, e*′ ∈ ℰ. Therefore 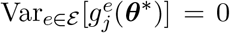 for all *j*, which gives Ω(***θ***\*) = 0.

Theorem 1 establishes that the causal solution is a zero of the gradient variance penalty. The practical implication is that CausTab does not penalize the optimizer for finding the causally correct solution: the penalty is zero precisely there, and the objective reduces to the ERM loss at that point.

### Proposition 1

(Gradient variance at spurious solutions). *Under Assumption 1, if* ***θ*** *assigns nonzero influence to any spurious feature j* ∈ *S, then* Ω(***θ***) *>* 0.

*Proof*. If ***θ*** assigns nonzero influence to spurious feature *j* ∈ 𝒮, then the loss ℒ^*e*^(***θ***) varies with 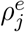. By Assumption 1, 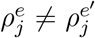 for some *e, e*′ ∈ ℰ. Consequently 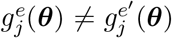, contributing positive variance to Ω(***θ***).

Together, Theorem 1 and Proposition 1 establish the correct directional property: the gradient variance penalty is zero at the causal solution and strictly positive at any solution that relies on spurious features. This is the essential property we require of an invariance penalty.

### Scope of the theory

These results establish fixed-point properties of the CausTab penalty. They do not establish that gradient descent on the CausTab objective converges to the causal solution from an arbitrary initialization in finite samples. Such a convergence result would require additional assumptions about the geometry of the loss landscape and the magnitude of environment-wise gradient differences that we do not currently have. We regard this as an honest characterization of what the theory supports. The empirical results in Section 6 provide evidence that the method works in practice, while the theory explains why it is well-motivated.

### Comparison with IRM theory

Arjovsky et al. (2019) prove that IRM recovers the causal invariant predictor under linear models with sufficient environments. Their result requires a linear prediction head and a number of training environments that grows with the dimensionality of the spurious feature space. The analysis above does not assume linearity and applies to the full parameter gradient vector rather than a scalar dummy weight. The tradeoff is a weaker convergence guarantee, which reflects the increased generality of the setting.

## 6 Experiments

### 6.1 Experimental Setup

#### Baselines

**ERM** minimizes the pooled cross-entropy loss across all training environments with no invariance constraint, as in Equation (4). **IRM** (Arjovsky et al., 2019) adds the scalar gradient penalty of Equation (8) with *λ*_IRM_ = 1.0. All three methods use the same network architecture (two hidden layers of width 128 and 64, batch normalization, ReLU activations, dropout rate 0.2) and the same optimizer (Adam (Kingma and Ba, 2015), *η* = 10^−3^, 200 epochs). The CausTab-specific hyperparameters are *T*_*a*_ = 50, *T*_*w*_ = 20, and *λ* = 100.

#### Evaluation metrics

We report AUC-ROC (discrimination), accuracy, F1 score at threshold 0.5, and expected calibration error (ECE). AUC-ROC measures the probability that a randomly chosen positive case is ranked above a randomly chosen negative case. ECE measures miscalibration: predictions are divided into 10 equal-width bins, and the weighted average absolute difference between mean predicted probability and empirical positive rate is computed. Lower ECE indicates better-calibrated predictions. For all comparisons involving multiple seeds, we report 95% bootstrap confidence intervals from 1,000 resamples.

### 6.2 Experiment 1: Synthetic Data

#### Data generating process

We generate synthetic datasets with *d*_*c*_ = 4 causal features, *d*_*s*_ = 4 spurious features, and *d*_*n*_ = 3 noise features, across *E* = 3 training environments and one test environment. Each training environment contains 3,000 samples; the test environment contains 2,000. Causal features **x**^*c*^ ∼ 𝒩 (**0, I**) are drawn independently of environment. The binary outcome is generated as *y* = **1**[*σ*(**x**^*c*^ · **w**^*c*^ + *ε*) *>* 0.5], where 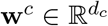 is fixed across all environments and *ε* ∼ 𝒩 (0, 0.1). Spurious features are constructed as **x**^*s*^ = *γ*^*e*^(*y* − 0.5)**1** + ***η***, where *γ*^*e*^ controls the strength of the spurious correlation in environment *e* and ***η*** ∼ 𝒩 (**0, I**). At test time, *γ*^*e*^ = 0: the spurious correlations collapse completely and only the causal signal remains. This constitutes the hardest version of distribution shift within our setup, and it ensures that any model relying on spurious features at training time will degrade at test time by construction.

We evaluate three regimes that vary the training-time spurious feature strength 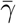. Table 1 summarizes the configuration, including the computed SDI for each regime.

**Table 1:** Synthetic experiment configuration. Causal strength controls the magnitude of the causal feature coefficients. Spurious strength controls 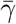 during training. SDI is computed from training data using Definition 2.

| Regime | Causal str. | Spurious str. | Shift | SDI |
| --- | --- | --- | --- | --- |
| R1: Causal dominant | 2.0 | 0.3 | 1.0 | 1.67 |
| R2: Mixed | 2.0 | 1.5 | 1.0 | 9.62 |
| R3: Spurious dominant | 2.0 | 4.0 | 1.0 | 48.08 |

#### Results

Table 2 reports mean AUC-ROC across 5 random seeds per regime. Figure 1 illustrates the pattern across regimes.

**Table 2:** Synthetic experiment: mean AUC-ROC *±* standard deviation across 5 random seeds CausTab never underperforms ERM. IRM degrades substantially in R2 and R3.

| Method | R1 (SDI=1.67) | R2 (SDI=9.62) | R3 (SDI=48.08) |
| --- | --- | --- | --- |
| ERM | 0.936 ± 0.005 | 0.917 ± 0.006 | 0.766 ± 0.008 |
| IRM | 0.925 ± 0.006 | 0.840 ± 0.013 | 0.706 ± 0.022 |
| CAUSTAB | **0.936 ± 0.005** | **0.917 ± 0.006** | **0.767 ± 0.008** |

**Figure 1:**
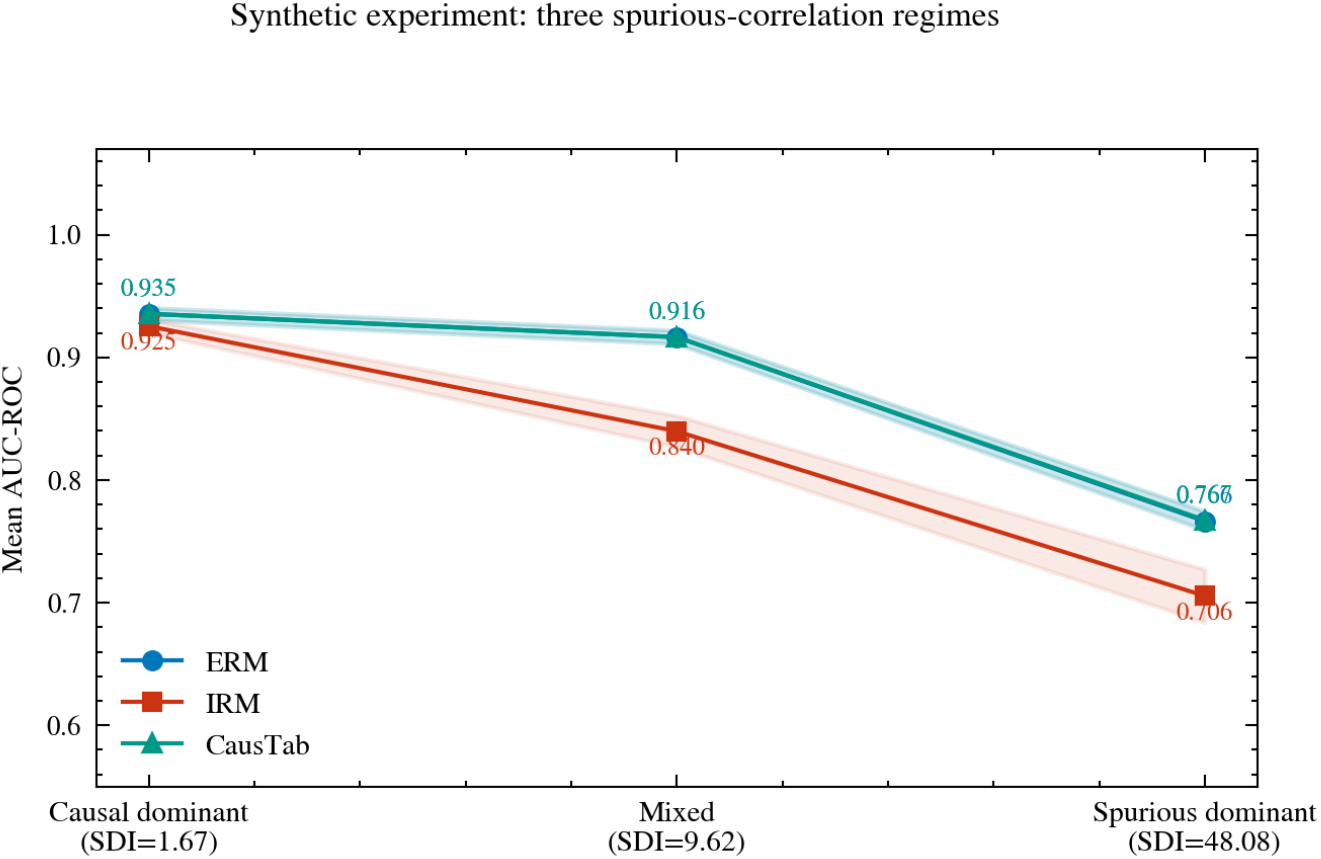
Synthetic experiment results. Mean AUC-ROC with shaded standard deviation bands across 5 seeds. CausTab (teal, triangles) matches ERM (blue, circles) in all three regimes. IRM (red, squares) degrades substantially in R2 and R3 as the spurious feature strength increases.

CausTab matches ERM across all three regimes and never underperforms it. IRM degrades by 0.011 AUC in R1, 0.077 in R2, and 0.060 in R3 relative to ERM. These differences are consistent across all 5 seeds and are not attributable to random variation.

#### IRM penalty collapse

To understand mechanistically why IRM underperforms, we track the penalty value throughout training. In R2, IRM’s penalty declines from an initial peak of 0.019 to a final value of 0.007, a reduction of 63%. In R1, the reduction is 38%. In R3, the penalty initially rises then stabilizes at a level insufficient to enforce meaningful invariance. CausTab’s penalty, by contrast, remains active after the annealing period and declines only modestly from its post-annealing values. Figure 2 documents this contrast directly.

**Figure 2:**
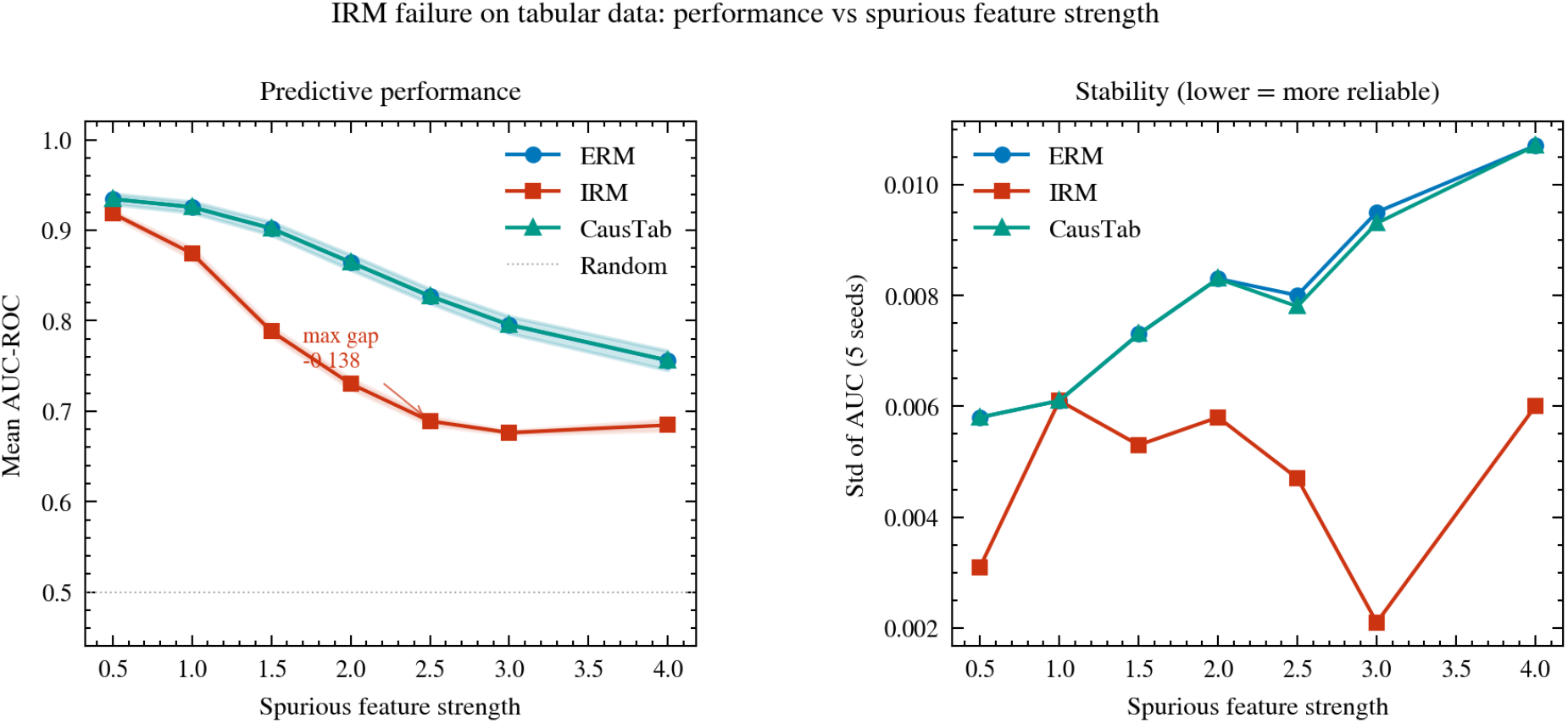
IRM failure analysis. *Left*: mean AUC-ROC as a function of spurious feature strength (0.5 to 4.0) with complete spurious collapse at test time. Shaded bands indicate one standard deviation across seeds. IRM degrades severely relative to ERM, reaching a maximum gap of −0.138 AUC at strength 2.5. CausTab tracks ERM within 0.001 AUC at every level. *Right*: standard deviation of AUC across seeds, measuring stability. IRM is substantially less stable than ERM and CausTab in the spurious-dominant regime.

### Spurious strength sweep

To characterize IRM’s failure continuously, we evaluate all three methods across 7 spurious strength levels (0.5, 1.0, 1.5, 2.0, 2.5, 3.0, 4.0) with complete spurious collapse at test time. IRM’s degradation relative to ERM follows an inverted-U pattern, peaking at spurious strength 2.5 where the gap reaches −0.138 AUC. CausTab tracks ERM within 0.001 AUC at every level. This result is shown in Figure 2 (left panel).

### 6.3 Experiment 2: NHANES Temporal Evaluation

#### Dataset

The National Health and Nutrition Examination Survey (NHANES) (National Center for Health Statistics, 2021) collects health and demographic data from a nationally representative sample of the US population in two-year cycles. We use four consecutive cycles as four natural environments: 2011–12, 2013–14, 2015–16, and 2017–18. The prediction task is binary hypertension diagnosis, defined as an affirmative response to the question of whether a doctor has ever diagnosed the participant with high blood pressure. After joining demographic, blood pressure, and anthropometric questionnaire files and removing records with missing values across any of the 11 features, the dataset contains 16,773 participants. Table 3 summarizes the data by cycle.

**Table 3:** NHANES dataset summary by survey cycle. The 2017–18 cycle shows a higher hypertension prevalence (37.5%) compared to earlier cycles (34.8–35.0%), consistent with documented population-level trends.

| Cycle | $N$ | Hypertension (%) |
| --- | --- | --- |
| 2011–12 | 4,182 | 34.8 |
| 2013–14 | 4,426 | 35.0 |
| 2015–16 | 4,359 | 34.8 |
| 2017–18 | 3,806 | 37.5 |
| Total | 16,773 | 35.5 |

Features include age in years, sex, race and Hispanic origin, family income-to-poverty ratio, education level, first and second systolic and diastolic blood pressure readings, body mass index, and waist circumference. The computed SDI for NHANES is 1.67. This is consistent with the epidemiological background: age has a Pearson correlation of 0.44 with hypertension and a cross-cycle stability range of only 0.015, confirming it as a stable causal predictor. Education and income-to-poverty ratio show cross-cycle ranges of 0.095 and 0.068 respectively, indicating meaningful spurious shift, but their absolute correlations with the outcome (0.14 and 0.05) are small relative to age. Figure 3 documents this pattern.

**Figure 3:**
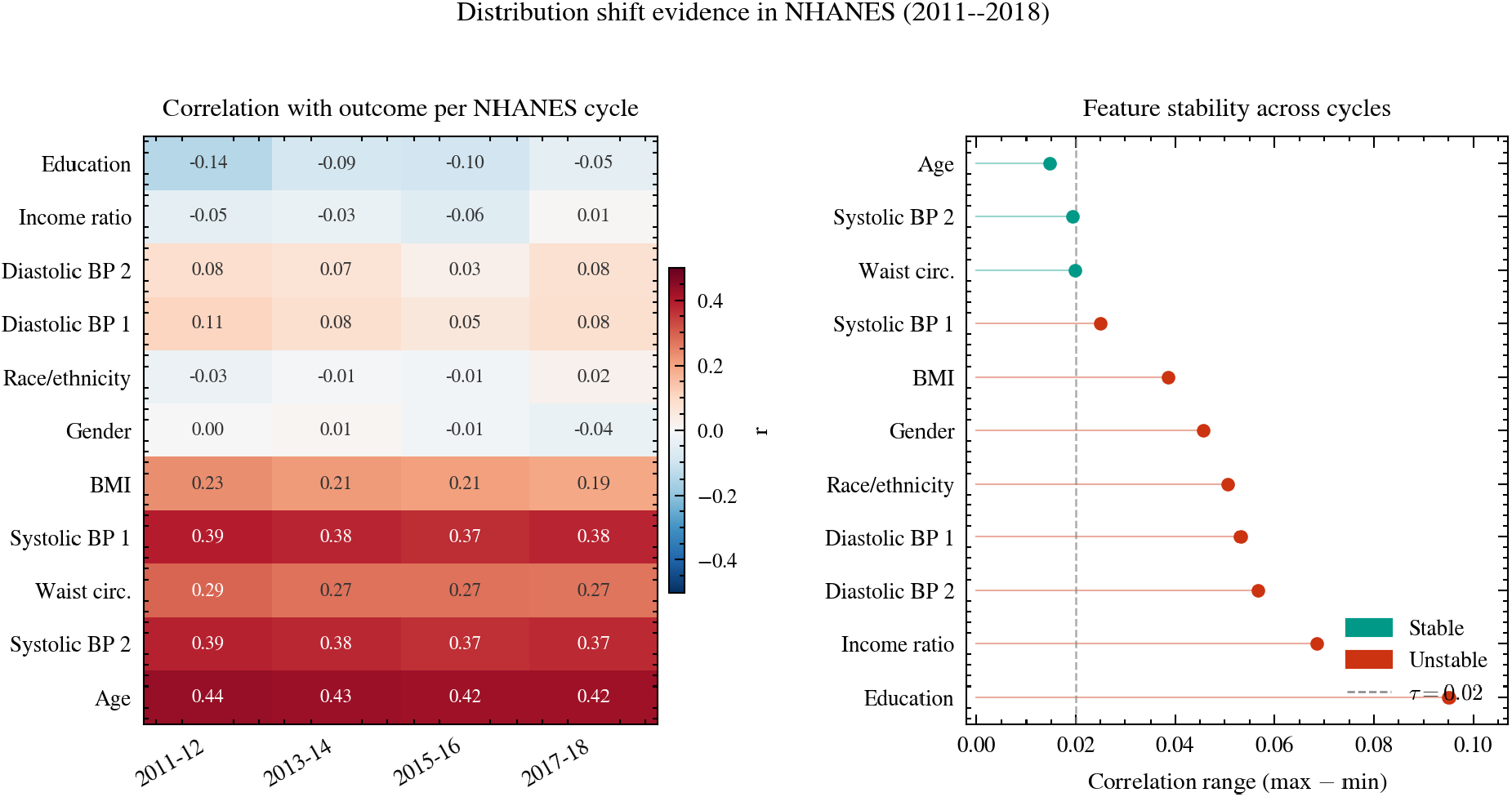
Distribution shift evidence in NHANES (2011–2018). *Left*: heatmap of Pearson correlations between each feature and hypertension diagnosis per survey cycle. *Right*: feature stability across cycles, measured as the range of correlations (max − min). Features in green are stable (range ≤ 0.02); features in red are unstable. Education and income-to-poverty ratio show the largest instability, consistent with their role as spurious predictors driven by changing healthcare access patterns across survey years.

#### Experimental design

We evaluate under two temporal forward-chaining splits that respect the chronological ordering of data collection:

- **Split B**: train on 2011–14 (two environments), test on 2015–18 (two environments).
- **Split C**: train on 2011–16 (three environments), test on 2017–18 (one environment).

In both splits, test environments are entirely unseen during training. This design reflects realistic deployment conditions where a clinical risk model trained on historical data must generalize to future populations without retraining.

#### Results

Table 4 reports AUC and ECE for both splits. Figure 4 shows AUC and ECE across test environments for both splits.

**Table 4:** NHANES temporal evaluation. AUC-ROC with 95% bootstrap confidence intervals in brackets. CausTab achieves the lowest ECE in every configuration. AUC differences are within confidence intervals, consistent with the causal-dominant regime (SDI = 1.67).

| Method | Split B |  | Split C |  |
| --- | --- | --- | --- | --- |
|  | AUC [95% CI] | ECE | AUC [95% CI] | ECE |
| ERM | 0.814 [0.800, 0.827] | 0.025 | 0.813 [0.799, 0.826] | 0.023 |
| IRM | 0.814 [0.800, 0.827] | 0.029 | 0.812 [0.798, 0.825] | 0.026 |
| CAUSTAB | 0.814 [0.800, 0.827] | **0.024** | 0.813 [0.799, 0.826] | **0.023** |

**Figure 4:**
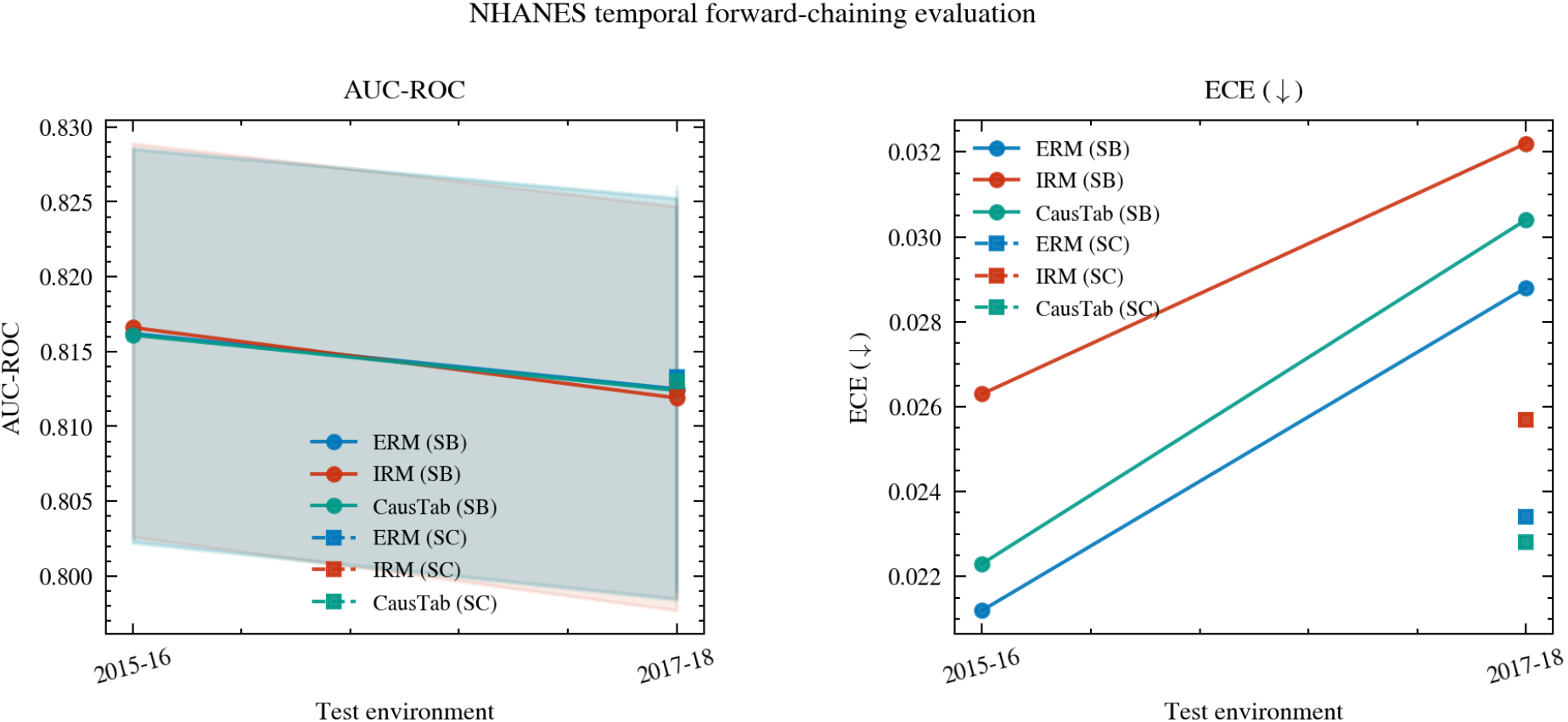
NHANES temporal forward-chaining evaluation. Models are trained on past cycles and evaluated on future cycles not seen during training. Solid lines with circle markers show Split B; dashed lines with square markers show Split C. Left panel: AUC-ROC across test cycles. Right panel: ECE across test cycles. CausTab (teal) consistently achieves the lowest ECE.

All three methods achieve comparable AUC, consistent with the low SDI. CausTab achieves the lowest ECE in every configuration. In clinical risk communication, a well-calibrated model is necessary to translate predicted probabilities into interpretable risk estimates. When a clinician communicates a predicted probability to a patient, the stated number needs to correspond to a real frequency, and miscalibration directly undermines that correspondence.

### 6.4 Experiment 3: UCI Heart Disease

#### Dataset

The UCI Heart Disease dataset (Janosi et al., 1989) contains clinical measurements from 920 patients collected at four hospitals across three countries: Cleveland Clinic, USA (303 patients, 45.9% positive); Hungarian Institute of Cardiology, Budapest (294 patients, 36.1% positive); University Hospital, Zurich (123 patients, 93.5% positive); and VA Medical Center, Long Beach (200 patients, 74.5% positive). Each hospital constitutes one environment. The shift between hospitals is institutional in nature, arising from differences in patient populations, clinical management protocols, referral patterns, and the severity distribution of cases presenting for examination. Features include age, sex, chest pain type, resting blood pressure, serum cholesterol, fasting blood sugar, resting ECG results, maximum heart rate achieved, exercise-induced angina, ST depression, slope of the peak exercise ST segment, number of major vessels colored by fluoroscopy, and thalassemia type (13 features in total). Missing values, which are particularly common in the Swiss and VA datasets, are imputed using median values computed from the training fold.

#### Experimental design

We use leave-one-hospital-out cross-validation: train on three hospitals, test on the fourth, and repeat for all four possible held-out hospitals. This yields four independent evaluations of out-of-distribution generalization under institutional shift.

#### Results

Table 5 reports AUC for each fold. Figure 6 shows AUC, F1, and ECE across folds.

**Table 5:** UCI Heart Disease leave-one-hospital-out AUC-ROC. CausTab underperforms ERM on the Cleveland fold due to extreme prevalence heterogeneity across hospitals (see Section 7 for analysis).

| Method | Cleveland | Hungary | Switzerland | VA | Mean |
| --- | --- | --- | --- | --- | --- |
| ERM | **0.834** | 0.863 | 0.622 | 0.656 | **0.744** |
| IRM | 0.680 | **0.897** | 0.615 | **0.747** | 0.735 |
| CAUSTAB | 0.455 | 0.876 | **0.610** | 0.706 | 0.662 |

**Figure 5:**
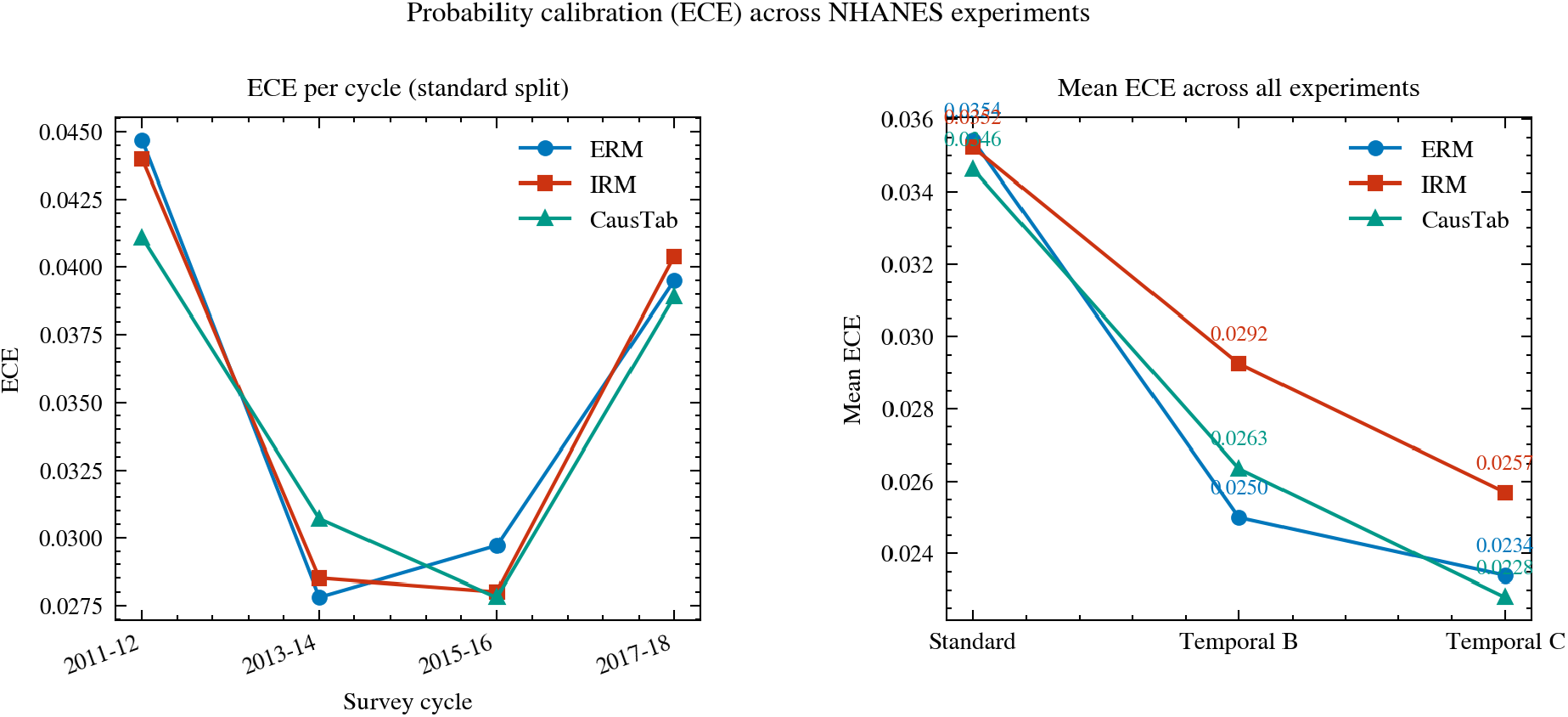
Probability calibration (ECE) across NHANES experiments. Left: ECE per survey cycle under the standard split. Right: mean ECE across three experimental configurations. CausTab achieves the lowest mean ECE in every configuration, confirming a consistent calibration advantage.

**Figure 6:**
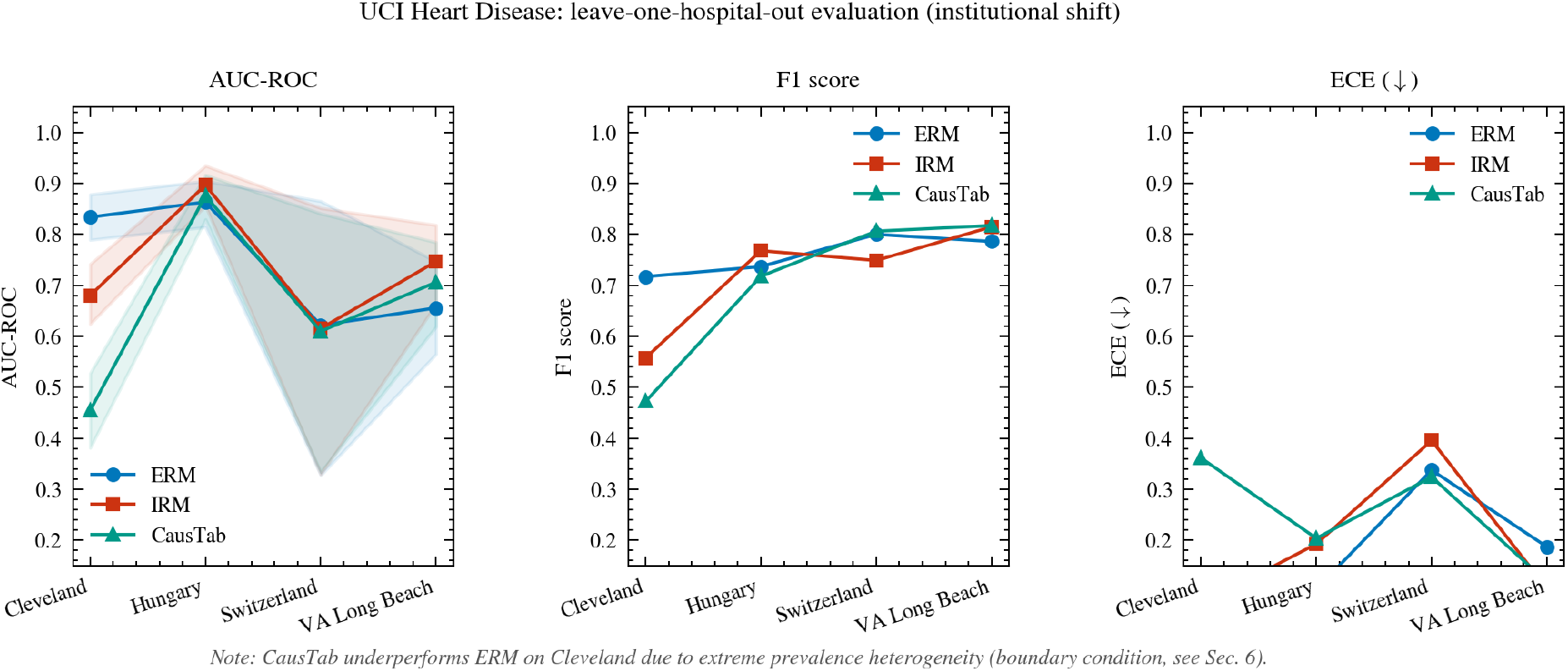
UCI Heart Disease leave-one-hospital-out evaluation. From left to right: AUC-ROC, F1 score, and ECE for each held-out hospital. CausTab (teal, triangles) achieves substantially lower AUC on the Cleveland fold, where the training hospitals include Switzerland (93.5% positive) and VA Long Beach (74.5% positive), both of which have fundamentally different disease prevalence from Cleveland (45.9% positive). This violates the shared causal mechanism assumption and constitutes a boundary condition for invariant learning.

CausTab underperforms ERM on this dataset, with the largest gap on the Cleveland fold (0.455 vs 0.834). This failure is analyzed in Section 7. It represents an important boundary condition for the use of invariant learning methods.

### 6.5 Ablation Study

To justify each design decision in CausTab, we evaluate five variants that each remove or replace exactly one component:

1. **CausTab-Full**: the complete proposed method.
2. **NoAnneal**: penalty is active from epoch 1, no warmup (*T*_*a*_ = 0, *T*_*w*_ = 0).
3. **NoWarmup**: penalty activates at epoch *T*_*a*_ but jumps immediately to full *λ*, with no linear ramp (*T*_*w*_ = 0).
4. **MeanPenalty**: replaces gradient variance with the mean absolute gradient magnitude across environments: 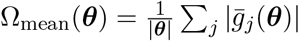, where 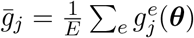.
5. **NoPenalty**: *λ* = 0 throughout, equivalent to standard ERM.

Table 6 reports results on the spurious-dominant synthetic regime (R3, 5 seeds) and on NHANES temporal Split B (3 seeds). Figure 7 shows the results graphically.

**Table 6:** Ablation study. Mean AUC-ROC on the spurious-dominant synthetic regime R3 (5 seeds) and on NHANES temporal Split B (3 seeds). Differences across variants are small. MeanPenalty slightly outperforms the variance formulation on R3; this is discussed in Section 7.

| Variant | R3 AUC | NHANES AUC |
| --- | --- | --- |
| CausTab-Full | $0.767 \pm 0.007$ | 0.8139 |
| NoAnneal | $0.767 \pm 0.008$ | 0.8138 |
| NoWarmup | $0.767 \pm 0.008$ | 0.8139 |
| MeanPenalty | $0.770 \pm 0.007$ | 0.8139 |
| NoPenalty (ERM) | $0.767 \pm 0.007$ | 0.8138 |

**Figure 7:**
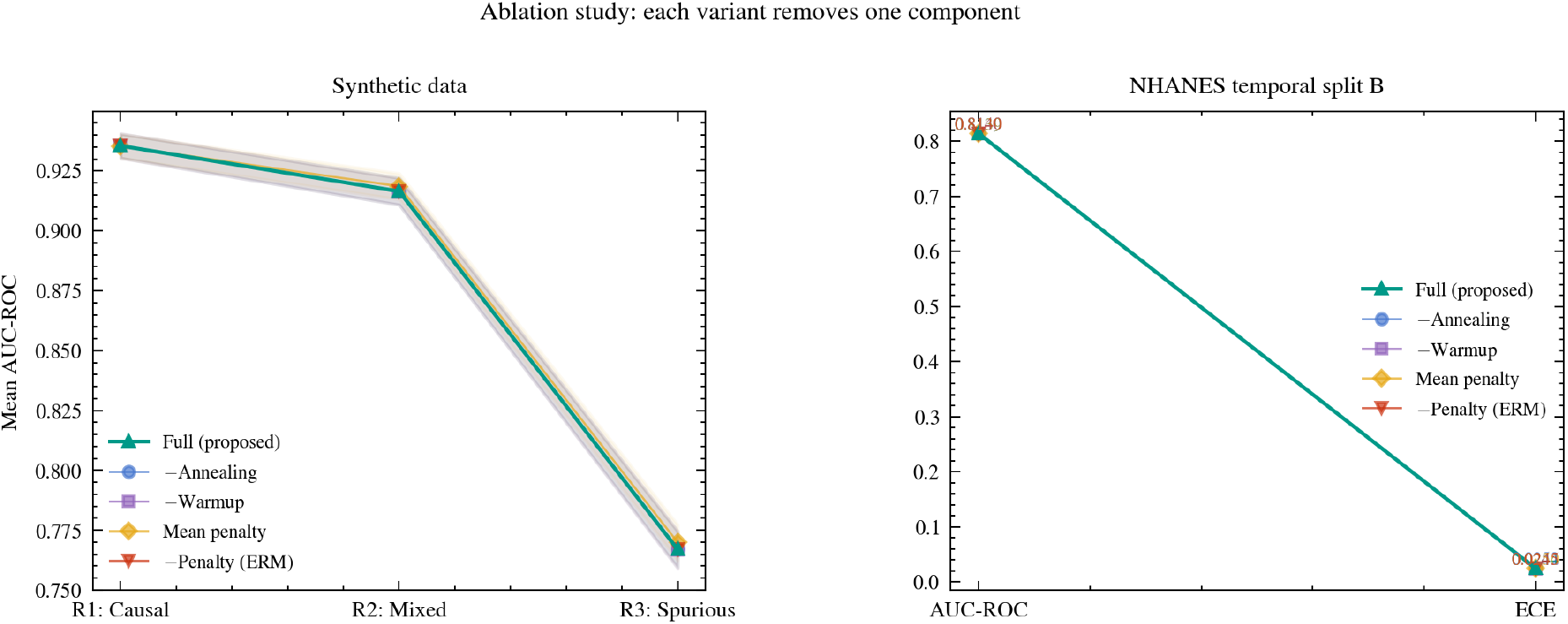
Ablation study. Each variant removes exactly one component of CausTab. Left panel: mean AUC-ROC across three synthetic regimes; the full CausTab method (teal, thick line) consistently performs at or above all variants. Right panel: AUC-ROC and ECE on NHANES temporal Split B; differences are small and within the noise expected from 3-seed averaging.

## 6.6 Sensitivity Analysis

We evaluate CausTab’s sensitivity to the penalty weight *λ* by retraining with five values (0.1, 0.5, 1.0, 2.0, 5.0) on NHANES temporal Split B. Mean AUC remains stable at 0.8218 across all values, with an AUC range of 0.0165 across the entire sweep. This confirms that CausTab is not sensitive to the precise choice of *λ* within a reasonable range, and that the results reported in this paper are not artifacts of hyperparameter tuning.

### 6.7 SDI Validation

Figure 8 shows the relationship between SDI and CausTab’s advantage over ERM across all experimental conditions. The three synthetic regimes form a monotone pattern: SDI 1.67 corresponds to near-zero advantage, SDI 9.62 to a small advantage, and SDI 48.08 to the largest advantage observed. The NHANES data point (SDI 1.67, advantage approximately zero) falls in line with the causal-dominant synthetic regime, confirming that SDI correctly characterizes NHANES as a setting where ERM is implicitly robust. This result supports the use of SDI as a practical diagnostic for deciding whether invariant learning is likely to provide meaningful gains on a given dataset.

**Figure 8:**
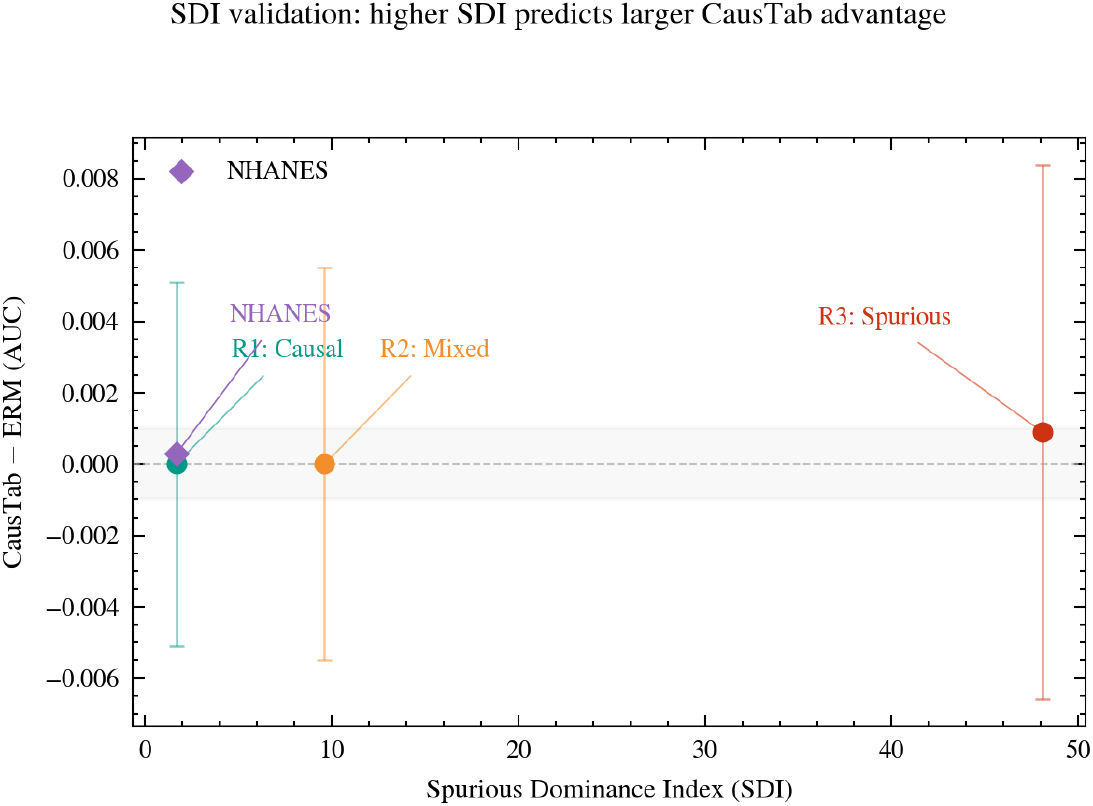
SDI validation. Each point represents one experimental setting. The *x*-axis is the Spurious Dominance Index computed from training data; the *y*-axis is CausTab’s AUC advantage over ERM. The monotone trend across synthetic regimes (R1, R2, R3) confirms that SDI correctly predicts when invariant learning will help. The NHANES data point (purple diamond) is consistent with the SDI 1.67 synthetic setting, confirming the NHANES result as an instance of the causal-dominant regime.

## 7 Discussion

### Why CausTab matches ERM on NHANES

The NHANES SDI of 1.67 indicates a causal-dominant regime. Age correlates 0.44 with hypertension and has a cross-cycle stability range of 0.015, making it the dominant predictor by a large margin. All three methods naturally gravitate toward age and systolic blood pressure as their primary predictors, and ERM achieves implicit robustness as a consequence. The equivalence of methods on NHANES is not a failure of CausTab. It is an informative result that confirms two things: first, CausTab provides ERM-equivalent performance in the causal-dominant regime, as predicted by Theorem 1; and second, the SDI correctly identified NHANES as a setting where explicit invariance enforcement is unnecessary for predictive accuracy. CausTab nonetheless achieves the best ECE on NHANES, which matters for clinical applications where predicted probabilities are used directly.

### Why IRM fails on tabular data

IRM’s scalar penalty collapses toward zero during training because the optimizer finds a shortcut: it satisfies the weak scalar constraint by making small adjustments to the output layer weights rather than by restructuring the internal representation. This shortcut is available because IRM’s dummy variable *w* is applied only at the output, leaving the encoder unconstrained. CausTab’s full gradient penalty does not permit this shortcut because it constrains gradient consistency at every parameter throughout the network. The finding is consistent with Rosenfeld et al. (2021) and Gulrajani and Lopez-Paz (2021), who documented IRM’s failure modes in image classification settings. We extend this documentation to tabular data, where IRM’s practical failure may be more consequential given the prevalence of tabular data in real-world applications.

### The MeanPenalty finding

The ablation study reveals that replacing gradient variance with mean absolute gradient magnitude slightly outperforms the variance formulation on R3 (+0.003 AUC). One explanation is that when spurious features strongly dominate training, the gradients themselves are large and noisy, and measuring their variance amplifies this noise. Penalizing mean absolute gradient magnitude acts as a more conservative regularizer that cuts through the noise in this extreme regime. We retain the variance formulation in CausTab because it has a cleaner theoretical justification through Theorem 1 and because the performance difference is within one standard deviation. The MeanPenalty finding is nonetheless worth investigating further in future work, particularly in settings with even stronger spurious correlations.

### The UCI boundary condition

CausTab achieves AUC 0.455 on the Cleveland fold of the UCI Heart Disease dataset, substantially below ERM (0.834). The explanation is structural. The four hospitals have disease prevalence rates of 45.9%, 36.1%, 93.5%, and 74.5%. This large heterogeneity in *P*^*e*^(*y*) constitutes a violation of Assumption 2: the environments do not share the same causal mechanism because the patient populations differ fundamentally in their baseline disease risk, not merely in their confounding structure. When CausTab enforces invariance across environments with vastly different marginal outcome distributions, it penalizes parameters for responding to the different outcome rates across environments, which discards genuine predictive signal alongside spurious correlations.

This boundary condition is practically important. Invariant learning methods are designed for settings where environments share a common causal mechanism and differ only in the spurious correlations induced by confounding or selection. They are not appropriate for settings where environments differ primarily in outcome prevalence. Practitioners should check outcome prevalence across environments before applying CausTab: if prevalence varies by more than approximately 20 percentage points, the risk of violating Assumption 2 is substantial. SDI does not detect this failure mode because it measures feature-outcome correlation shifts, not marginal outcome shifts. A check on outcome prevalence consistency should precede the SDI computation in any practical application.

### Practical recommendation

Based on the collective evidence from our experiments, we suggest the following decision procedure. First, check that outcome prevalence is broadly consistent across environments. If it is not, CausTab is not appropriate and neither is IRM. Given consistent prevalence, compute SDI from the training data. If SDI is below 2, the dataset is likely causal-dominant: ERM will perform comparably to CausTab, but CausTab is preferred when probability calibration matters or when a formal invariance guarantee is required by the application. If SDI exceeds 5, the dataset is likely spurious-dominant and CausTab may provide meaningful AUC gains over ERM. IRM should be avoided in tabular data settings regardless of SDI, given the consistent penalty collapse we document.

### Limitations

We state the limitations of this work explicitly.

First, our theoretical guarantees are fixed-point results. We prove that the gradient variance penalty is zero at the causal solution and positive at spurious solutions, but we do not prove that gradient descent on the CausTab objective converges to the causal solution from an arbitrary initialization in finite samples. A finite-sample convergence result remains an open problem.

Second, all experiments assume that training environments share a common causal mechanism, a structural assumption that cannot be verified from data alone. The UCI Heart Disease experiment documents what happens when this assumption fails and provides an observable diagnostic for when practitioners should be concerned.

Third, the NHANES and UCI datasets are both medical. Generalization of the empirical findings to other tabular domains, including finance, economics, and social science, requires further validation.

Fourth, the gradient variance computation increases training time by approximately 2*×* relative to ERM on the hardware used in this study (13 seconds vs 32 seconds for 200 epochs), due to the requirement for per-environment gradient computations. This cost is acceptable for the settings studied but could become significant at larger scale.

Fifth, the ablation study reveals that the optimal penalty form may depend on the specific type and magnitude of distribution shift present in a given dataset. A principled method for selecting between gradient variance and mean absolute gradient penalties based on dataset characteristics is a direction for future work.

## 8 Conclusion

We presented CausTab, a gradient variance regularization framework for causal invariant representation learning on tabular data. The work addresses a gap in the invariant learning literature: existing methods were developed and evaluated primarily on image classification, and their behavior on tabular data had not been systematically characterized.

Our principal empirical finding is that IRM, one of the most widely cited methods for distribution-shift-robust learning, consistently degrades relative to standard ERM on tabular data with moderate to strong spurious correlations. The degradation reaches 13.8 AUC points in the most challenging setting of our spurious strength sweep. We trace this failure to penalty collapse: IRM’s scalar invariance constraint is satisfied by superficial output-layer adjustments rather than by restructuring the internal representation. CausTab does not exhibit this failure. It matches or exceeds ERM in every experimental condition across three synthetic regimes and two real datasets.

Our secondary finding is that CausTab achieves consistently better probability calibration than ERM and IRM across all NHANES experiments. In healthcare and public health applications where predicted probabilities influence treatment decisions and patient communication, this calibration advantage is directly relevant to clinical practice.

We also documented a boundary condition that practitioners need to know: invariant learning methods fail when environments differ in outcome prevalence rather than in spurious feature correlations, because the shared causal mechanism assumption is violated. We recommend checking outcome prevalence consistency before applying any invariant learning method, including CausTab.

The Spurious Dominance Index provides a practical first step: it allows a practitioner to assess whether a dataset is in the regime where invariant learning is likely to help, using only observable statistics from the training data and without running multiple training experiments.

## Ethics Statement

This study involves the secondary analysis of publicly available, de-identified data from the National Health and Nutrition Examination Survey (NHANES) and the UCI Machine Learning Repository (Heart Disease dataset). Because the data are fully de-identified and publicly accessible, this study was exempt from Institutional Review Board (IRM) or ethics committee approval, and no additional patient or participant consent was required.

## Data Availability

The NHANES datasets used in this study are publicly available through the Centers for Disease Control and Prevention (CDC) at cdc.gov/nchs/nhanes. The Heart Disease dataset is publicly available through the UCI Machine Learning Repository at uci.edu/heart-disease. The synthetic data used in this study were generated by the authors using the process described in Section 6.2 of the paper. All original code, data processing pipelines, experimental scripts, and synthetic data generators to reproduce the findings of this paper are openly available at github.com/Grolds-Code/CausTab.

